# Impact of persistent COVID-19 symptoms on social life of female long haulers: A qualitative study

**DOI:** 10.1101/2022.01.21.22269671

**Authors:** Atefeh Aghaei, Ran Zhang, Slone Taylor, Cheuk-Chi Tam, Chih-Hsiang Yang, Xiaoming Li, Shan Qiao

## Abstract

**Introduction:** Persistent COVID-19 symptoms (long COVID) may bring numerous challenges to long haulers’ social lives. Women may have to endure more profound impacts given their social roles and existing structural inequality. This study aims to explore the impacts of long COVID on various aspects of social life among female long haulers.

**Methods:** We conducted 15 semi-structured interviews with female long haulers in the United States purposely recruited from Facebook groups, Slack groups, and organization websites. The interviews were audio recorded after appropriate consent and transcribed verbatim. Inductive approach was applied in thematic analysis, which consists of six stages: becoming familiar with data, developing initial codes, extracting themes, refining themes, labeling themes, and reporting. The MAXQDA software was used in data analysis.

**Results:** Persistent COVID-19 symptoms negatively affected female long haulers’ social lives in many aspects including physical function, financial security, social relationship, conflict of social roles, and social stigma. Physical limitations changed their body image. Social isolation and work-family conflicts caused huge stress. They experienced internalization of stigma and job insecurities. Shifting to new methods of communication, especially social media may buffer the negative effects of social isolation because of long COVID.

**Conclusion:** Existing policies and intervention programs need to be adapted to address the challenges and barriers that long haulers face in returning to normal social life, especially for females. Tailored social life-related recommendations and social support are needed for female long haulers.

## INTRODUCTION

The coronavirus disease 2019 (COVID-19) pandemic has resulted in 290 million infection cases globally with over 5.4 million deaths (1). Although many people infected with COVID-19 have no or mild symptoms that last about two weeks, some individuals experience persisting or cyclical symptoms even after recovering from the acute phase of the illness (2). According to the Centers for Disease Control and Prevention (CDC), the persistent symptoms related to COVID-19 can be multi-faceted, and the most common symptoms are physiological problems (e.g., fatigue and shortness of breath), while other problems include cognitive problems (e.g., difficulty concentrating), and emotional distress (e.g., depression) (3). The National Institutes of Health (NIH) refers to such long-term COVID-19 symptoms as post-acute sequelae of SARS-CoV-2 (PASC), more commonly referred to as “post-COVID syndrome”, “long COVID”, or “long-term COVID” (4-6). People with post-COVID syndrome are referred to as “long haulers” (7). These persistent symptoms can present as different types and combinations of health problems over different time periods, thus significantly affecting many aspects of people’s social lives (e.g., interactions, relations) (8).

Notably, female long haulers are disproportionately affected by long COVID. The U.S. Department of Health and Human Services (9) reports that women comprise up to 80% of the patient population suffering from persistent symptoms. Although preliminary reports indicate that women have a reduced risk of severe disease and death from COVID-19 compared to men (10, 11), the discussion of the impact of long COVID effects on social life should account for unique characteristics that make women vulnerable to conditions that affect their health status (12). Furthermore, women face specific social stressors due to the pandemic or long COVID, including social isolation, unstable income, and the burden of caring for their families (10). The combination and interaction of these stressors can negatively impact the social aspects of the lives of female long haulers.

Health challenges and inequalities faced by women during COVID-19 have been rooted in their social roles. Social roles are the basis of people’s sense of self, and each person establish his or her identity and behave according to their social roles (13). Meanwhile, as the surrounding environment changes, people’s social roles would change, and the change would lead to psychosocial distress without a rapid adjustment. In the context of COVID-19, collective efforts to end the pandemic have resulted in an unprecedented disruption of social systems and roles (14). For example, individuals who play social roles as parents, employees, and friends during COVID-19 face different responsibilities (e.g., home-schooled children), behavioral routines (e.g., wearing a mask in the workplace), contexts of role execution (e.g., online socialization), and even complete suspension from work (e.g., leave of absence). The increased responsibility of caring for family members during a lockdown can lead to physical and emotional exhaustion, stress, and burnout (15). Furthermore, the collective actions taken by society to end COVID-19 and the social roles of women experiencing long COVID lack continuity with their real roles prior to the pandemic.

COVID-19 also caused a tremendous crisis for the economy. The spread of the COVID-19 virus disrupted international supply chains and forced workers to stay home as they were quarantined, sickened or on lockdown. Companies across all industries were forced to disrupt and downsize their operations, with massive job losses ensuing (16). A COVID-19 study reported that approximately 15% of U.S. respondents had already reduced their wages or working hours in March 2020, and 1.5% had already lost their jobs (17). Another study indicated that 40% of Americans experienced one or more COVID-19-related financial stressors by mid-April 2020 (18). Meanwhile, the pandemic magnified previously existing gender-based differences. Early studies have shown that women have been disproportionately affected by COVID-19-related unemployment and economic impacts compared to men (19, 20). During COVID-19, women’s lower status in the labor market leaves them exposed and more vulnerable to dismissal. Data from the KFF Women’s Health Survey showed that 8% of women reported quitting their jobs for COVID-19-related reasons (21). Among women who quit their jobs for COVID-19-related reasons, 48% said they quit because they felt unsafe in the workplace, 30% said it was because their children’s school or daycare center was closed, and nearly 20% said they quit because they lived with someone at higher risk of being infected. These impacts threaten to roll back already fragile gains in female labor force participation and limit women’s ability to support themselves and their families (22).

While the COVID-19 pandemic and persistent symptoms have affected people of all genders, women have faced many challenges due to their social roles and existing structural inequalities. However, there is a dearth of empirical studies that show their lived experiences and comprehensively explore the consequences of long COVID on their life. Therefore, the current qualitative study aims to characterize the impacts of long COVID on various aspects of social life among female long haulers.

## METHODS

### Study design

The current study was based on an online health promotion intervention among long haulers. The long haulers interviewed for this study were participants enrolled in the online health promotion intervention. Participants were recruited primarily using social media, with Facebook being the main social media platform through purposive and snowball sampling. Eligibility criteria included living in the United States, being 18 years of age or older, speaking and understanding English, having been infected with COVID-19, and having experienced at least one COVID-19 symptom lasting four weeks or longer after a COVID-19 diagnosis. There were 16 Facebook groups, one Slack group, and two organizations’ websites identified relating to these topics. Upon receiving approval from administrators of the groups and organizations, posts were made to eight Facebook groups, one Slack group, and one organization’s website, which included the study description and contact information for researchers, and a recruitment flyer attached to the posts. Recruitment took place over two and a half months from the end of March 2021 to mid-June 2021. In total, 17 participants completed a written informed consent and participated in a semi-structured interview. Fifteen of them qualified for the objectives of this study. This study was reviewed and approved by the University of South Carolina Institutional Review Board (Pro00109358).The semi-structured interviews were conducted online using the video teleconferencing software Zoom (23). Interviews were scheduled according to the schedule of the interviewer and participants, and each interview was held one-on-one with only the interviewer and each participant present. The interviews coincided with recruitment and took place from the beginning of April 2021 to the beginning of June 2021.

Table 1 shows the characteristics of the participants. Most participants were between the ages of 36 and 65 years (n=12). The primary distributions of participants by occupation were in the health care (n=5), education (n=4), and business field (n=4). Since social media and organization’s website were used for posting recruitment information, the geographic location of participants were concentrated in the eastern states (n=10).

**Table 1.** Demographic characteristics of female long haulers.

| **Variable** | **n (total=15)** | **Percent** |
| --- | --- | --- |
| **Age** |  |  |
| 20-35 | 2 | 13.33 |
| 36-50 | 6 | 40.00 |
| 51-65 | 6 | 40.00 |
| > 65 | 1 | 6.67 |
| **Occupation** |  |  |
| Health Care Provider | 5 | 33.33 |
| Educator | 4 | 26.67 |
| Business Owner | 4 | 26.67 |
| Student | 1 | 6.67 |
| Retired | 1 | 6.67 |
| **Living situation** |  |  |
| Live with others | 13 | 86.67 |
| Live alone | 2 | 13.33 |
| **Location** |  |  |
| East | 10 | 66.67 |
| Central | 3 | 20 |
| West | 2 | 13.33 |

### Data collection

The primary purpose of the semi-structured interviews was to gather information on the impact of COVID-19 persistent symptoms on different aspects of the social life among female long haulers. The interview guide covered the background of COVID-19 infection and related symptoms, the psychosocial impact of COVID-19, and social sources for support and resilience. (Table 2). Participants were given a $30 Amazon e-gift card as compensation for their time.

**Table 2.** Semi-structured interview guide.

| **Topic** | **Question** |
| --- | --- |
| **Background of COVID-19 infection and its physical related symptoms** | 1. When were you diagnosed with COVID-19?<br>2. What symptoms have you experienced since the infection?<br>3. Do you experience any long-term (or persistent) symptoms, which last for weeks or even months after recovery?<br>4. Do these symptoms affect your life? (Social, working, daily life, etc.)<br>5. What are the biggest challenges that the COVID-19 infection and long-COVID symptoms brought to you? |
| **Psychosocial influences of COVID-19** | 1. Did the symptoms effect your moods? Your mental health? (Describe some examples)<br>2. Do other people know that you have these symptoms?<br>3. How do they respond it? (Describe specific examples and/or situations) |
| **Social sources for support and resilience** | 1. Who do you turn to when you experience stress and distress? How did they help?<br>2. Do you need any additional resources or help to enhance your physical and mental health? If so, what are they?<br>3. Are there any things you do to help to overcome bad feelings and bounce up? |
| **Others** | 1. Do you have any questions for me about the study and intervention?<br>2. Is there anything else would you like to share with me? |

Field notes were completed after each interview by the interviewer. Each interview was recorded with verbal permission from the participant for transcription purposes. The transcription software Otter.ai (24)was used to transcribe the interviews, and they were reviewed and edited by the interviewer.

### Data analysis

The inductive approach for thematic analysis was applied in this study (25-27). The thematic analysis consists of six stages: becoming familiar with data, developing initial codes, extracting themes, refining themes, labeling themes, and reporting (28). The MAXQDA software (29) was used to analyze the interviewee transcripts. First, transcripts were reviewed multiple times for accuracy (30, 31), and emergent themes (open coding) were identified (32). Then, subthemes were categorized into five main themes (axial coding). In this thematic analysis, initial codes were discussed during team meetings. The final codebook was obtained after reorganizing the codes when required. This codebook contained definitions of the themes, exemplar quotes, and quote samples that did not fit into the categorization.

We then compared and contrasted themes to determine similarities, differences, and associations between findings. We employed standard and in-depth qualitative data analysis techniques during coding, including open coding, marginal remarks, axial coding, memo-writing, and comparisons. Peer debriefing and inter-coder agreement techniques were used to ensure the reliability of the analysis (33-35). Inter-coder agreement reliability is a measurement of agreement between the coders about the similar data (36). The codes and results were presented to two research team members who were not involved in the data analysis to discuss themes and related outcomes (33, 35). Verbatim quotes representing the themes were chosen to demonstrate the main findings.

## RESULTS

Persistent symptoms of female long haulers adversely affect their social lives in different ways. Interviews with female long haulers have identified five main categories: physical limitations, economic issues, social relationships, conflict of social roles, and social stigma. All main themes and their dimensions are illustrated in Fig.1.

**Figure 1.**
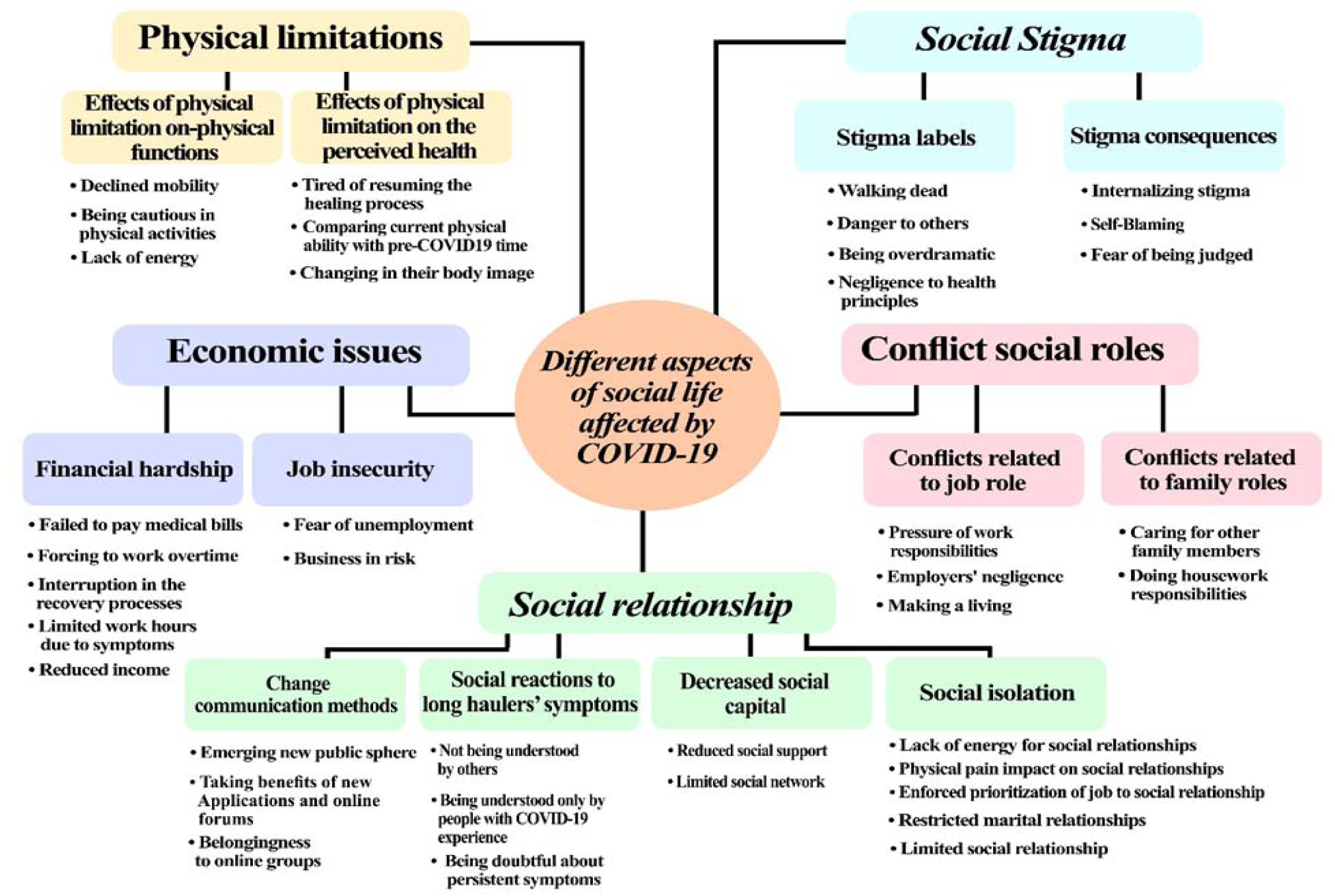
Dimensions of the COVID-19 persistent symptoms impact on social life of female long haulers.

### Physical limitation

Physical limitation is one of the issues that long haulers struggle with and affect their social life. The physical limitation has two dimensions: “Effects of physical limitation on the body” and “Effects of physical limitation on the mind.”

#### Effects of physical limitation on the physical function

Almost all the participants reported “declined mobility,” causing “*feelings of inactivity*.” For instance, even performing the simplest tasks, such as “*gardening or making a meal, “* led to negative feelings of being “*like a couch potato*.” Some participants reported that they had to” be cautious in physical activities*”* because they were still “*in and out of doctors’ offices*.*”* Almost all participants said they felt a “lack of energy” and quickly felt tired even after recovery. As a result, participating in social activities was difficult for them. One of them asserted, *“I can go out with a friend, but I can’t do anything after that*.*”*

#### Effects of physical limitation on the perceived health mind

Physical limitations also led to mental influence on female long haulers that even affected their recovery process and made it difficult for them to follow through with treatment. Some participants reported feeling “tired of resuming the healing process,*”* and they experienced feelings such as” *resentment”* toward the spirometer because they *“had to be still using it*.” More than half of the participants reported that at times they “compared current physical abilities with pre-COVID-19 time*”* and “*sighed for their current physical abilities*.” One participant said, *“this turned me into a homebody, which is anybody*.*”* Moreover, these physical limitations “changed their body image.” One participant said, “*all my muscle is gone and …mentally my body image had changed*.”

### Economic issues

The economic issue is another aspect of female long haulers’ social lives affected by COVID-19. For most participants, “*job insecurity*” and “*financial hardship*” were the main economic issues that impeded their recovery.

#### Job insecurity

Almost all the participants shared their concerns about their employment status. One participant reported the “*fear of being unemployment”* as even more stressful than the infection. Another participant said that a company *“can fire you for absolutely no reason*.” In this case, the participants were constantly looking for new jobs instead of focusing on their health improvement. One participant stated, *“they just push my resume aside, so that’s been more of a stressor to me than the pandemic*.*”* Two participants had to “*quit their jobs to attend to family-related affairs*.*”* Other participants noted that their *“*business is at risk*”* because of their persisting symptoms. One participant said, “*because I have to cook…I have to breathe*… *so my business took a hit*.*”*

#### Financial hardship

The impact of the pandemic on the domestic as well as the global economy has affected some participants’ businesses, resulting in a “reduction in their income.” A participant explained that she was “*forced to have a part-time job*.” However, many participants had to “*reduce their working hours*” *due to* persistent *symptoms*. One participant described the pain she felt while working. Another participant who works in the education system reported, “*I have lots of brain fog and completely forgetting what I was teaching*.” In addition, another participant stated, *“I’m being paid less because I’m not able to have full-time work*.” These issues contributed to the participants’ unstable financial status, thus to the point that they “failed to pay medical bills.” Some participants mentioned that they were concerned about medical costs due to the possibility of losing their job. One participant was worried that the government would *“stop paying COVID-19 claims*.*”* This situation prompted some participants to “*work overtime*.*”* In these circumstances, nearly half of the participants experienced “interruptions in the recovery processes*”* due to financial issues or lack of “*health insurance*.”

### Social relationship

Persistent symptoms affected the social relationships of female long haulers. The experiences shared by the participants can be classified into four themes: “social isolation,” “social reaction to long haulers’ symptoms,” “change communication methods,” and “decrease social capital.”

#### Social isolation

All participants experienced social isolation due to changes and “restrictions in social relationships.” These challenges were rooted in their “limited energy” and weal physical condition, making social interactions difficult. Several participants described the short time they spent with family and friends during the pandemic. One participant said, “*the biggest thing that has impacted me personally is the loss of socialization*.” In addition, some participants noted that staying at home and having limited relationships made them feel anxious and stressed. Some participants mentioned that kissing, hugging, and simply handshaking was done reluctantly to keep to manners. One participant said, “*I was free from the hospital, and my daughter finally could hug me*.” Several participants also mentioned some*” physical and emotional conflicts*” in dealing with social relationships. One participant said, “it’s *been hard jumping back into that social life*.*”* Additionally, another participant highlighted *“a change in personality”* by saying, *“I feel like I’m not as extroverted or bubbly as I used to be*.*”*

Persistent “physical pain also impacted the maintenance and development of their social relationships.” One participant said, “*if I’m on my feet too long*… *get pain*…*I haven’t been out with my friends*”. Many participants reported that COVID-19 changed and “restricted their marital relationships.” Some participants who were infected with their spouses were afraid to approach each other due to symptoms. Some participants described that they should “end part of their social relationships to save energy for work activities.” One of them said, *“if I go, then I’m probably gonna feel tired…I can’t risk that for my job”*.

#### Social reactions to long haulers’ symptoms

About half of the participants reported that some people, especially those who had never been infected, feel quite “doubtful about persistent symptoms.” One participant said, *“Some are incredulous that it’s happening. they are like, oh, just fight through it”*. Other participants mentioned, *“some medical professionals try to trivialize it or make their patients think that it perhaps might be in their head*.*”* Moreover, the situation of long haulers is not understandable for some people. One of the participants asserted, “*Many people don’t understand; I could only last for about 30 minutes*”. Several participants reported that “only people experienced COVID-19 could understand their feelings”, such as “*the long COVID-19 Facebook groups because they are also going through it*.*”*

#### Change communication methods

Most participants have experienced some forms of online communication. In this regard, some female long haulers said that they had already missed the opportunity to attend the public sphere. Still, through pandemic by the “emergence of new public spheres,” participants pointed out they “have taken benefit of different new applications and online forums” for social interaction culminated in the feeling of “belongingness.” One of the participants asserted, “*I liked that because you are not alone*.*”* Finally, as said by a participant about a long hauler advocacy group, *“they trying to fight for even like disability rights for us*.*”*

#### Decreased social capital

Conflict in COVID-19 vaccination beliefs caused a loss of social capital for a few female long haulers. Participants pinpointed that “reduced social support” and “limited their social network” are adverse outcomes of decreasing social capital. One of the participants highlighted her higher level of vulnerability and said, “I’m *a social person …but I’m more cautious now because of my symptoms, and I don’t like to be in crowds which I don’t know if people had their vaccine*”. The other one mentioned, “*I couldn’t really interact with people because there’s a lot of people that have not had their vaccine*.” Consequently, less than half of the participants decided to terminate their relationship as one of them mentioned, “I’ve *lost a lot of friends because of it. Because … they’re anti-vaccine*”. Another participant highlighted the roots of conflict in the beliefs to vaccination and said, “*suddenly, people that have never had a political or social opinion, now do and so* … *that’s how it’s really affected me*…”

### Conflicts of social roles

Most participants experienced conflicts in their social roles. Their primary role was being a patient, but as a patient, their performance in their social life had some conflicts with their ability. The conflicts in different social roles can be divided into two main themes: “conflicts related to job roles” and “conflicts related to family roles.”

#### Conflicts related to the job role

Almost all participants reported being employed prior to the pandemic. Their job roles expose them to different conflicts that revolve around their expectations and responsibilities in the workplace and their relationship with their employers.

Some participants were adversely affected by their” employer’s negligence*”*, which made them “*feel helpless and ignored*”. One of the participants said, “*They didn’t call us one time. I noticed that they weren’t really there for us”*. Some participants also reported not being permitted by their supervisors to leave the workplace when they had to or being unjustifiably pressured to work by their supervisors.

In terms of “making a living”, participants turned out to have been struggling with financial problems intensified by COVID-19. Since their employers did not support them, they were pushed to “work overtime” or to “*cover shifts when other staff members were infected*.” One participant said, *“when I was supposed to be on sick leave, I was in my bed, typing up stuff*.*”* In addition, people who work under such conditions experience fatigue; as a result, their poor performance at work leads to “*increased conflict with employers*.*”* Especially for the head of the household, work role conflicts can cause them to wrestle with unemployment that was even more demanding and stressful than COVID-19 infection. Thus, some participants felt they had no choice but to push themselves to challenge themselves to return to business. One participant said, “*I was trying to get to work and just struggling with that*.”

Some participants reported that “the pressure of work responsibilities” like safe workplace requirements during COVID-19 was inevitable. The requirement to wear a mask to work was very difficult for participants with breathing problems. One participant stated, “*professionally, that was the hardest thing, making sure that my students are safe*.” This situation has become even more complicated for health care providers. As one nurse said, “*I have never left my job because I have patients on my schedule*.*”*

#### Conflicts related to the family roles

Most participants mentioned that they were expected to take care of the family even when they struggled with their persistent symptoms. This situation elevates the risk of a painful managing task for female long haulers. One participant said, “*we both were positive…but I have to make sure my son is okay*”. Furthermore, participants mentioned more exceptions related to their unique role as mothers. One participant said, “*my son was so stressed because I couldn’t take him to practice*.*”* Additionally, “doing housework responsibilities” is another challenge for female long healers. Almost all the participants reported that it is “*difficult to handle daily house chores”* in addition to work-related issues throughout the pandemic. One participant said, “*even now, just cleaning my house sometimes is too much*.”

### Social stigma

Experiencing social stigma had a negative impact on the social life of female long haulers. Social stigma dimensions were categorized as “stigma labels” and “stigma consequences”

#### Stigma labels

All participants were labeled variously by other people. Two participants stated that they were identified as *negligent to health principles*, especially at the pandemic’s beginning. One of them acknowledged that people did not talk openly about being infected with COVID-19 due to fear of being stigmatized and blamed for*” their careless behavior that caused the infection*.*”* Another participant stated, “*we had this sense of what did we do wrong?*”. Moreover, another two participants mentioned that they were identified as “danger to others” and labeled “*risk factor*.*”* One participant stated, “*we felt like a leper*.*”* Others explained that their “*situation was unbelievable for some people”* and triggered another type of stigma like being “overdramatic*”* that was driven from a misapprehension of the COVID-19 deterioration. One of them noted, “*a lot of my family members got it, but they didn’t get the after symptoms…they think I’m maybe being overdramatic*.” Meanwhile, a few participants were labeled as “walking dead” people. One of them asserted, “there are *so many stories… they collapsed, and after an autopsy, you know, it’s in their lungs*.”

#### Stigma outcomes

Ultimately, Stigmatization caused participants to “internalize the stigma,” “self-blaming,” and “fear of being a judge.” Participants identified themselves through the labels perceived in their social lives. For example, one of the participants mentioned, “there’s *a lot of question about why me? What did I do to have?*” and “*you feel like you did something wrong*.*” another one explained she “blamed herself for not being serious*.” Also, less than half of the female long haulers in this study restricted their social relationships because they avoided facing judgment and stigma. One of the participants asserted, “*Any symptom that I have* … *it’s embarrassing because I have to turn down like going with my husband’s family* “. Other participants explained, “*I feel like I have to keep proving to people that* … *Stress of people getting believe it and not feel like I’m making this up*.”

## DISCUSSION

Regarding limited research in this area, this study is one of the first attempts to explore the experiences of female long haulers and the impact of COVID-19 on their social life. In our study, most of the participants reported limited physical activity and changes in body image. Besides, layoffs or increased family burdens due to the pandemic caused many financial problems for them. Although almost all participants asserted that their social relationships had changed profoundly, emerging different groups and novel methods of communication in social media provide them with opportunities to gain a sense of belonging and support. Our findings revealed that conflict in social roles was mainly related to the expectations of others without considering the persistent symptoms of women. This situation culminated in experiencing stigma and internalizing it, which adversely affected the social lives of female COVID-19 long haulers.

In our study, most of the participants mentioned that long-lasting symptoms of COVID-19 reduce their activity and mobility in different aspects of their daily lives. Additionally, our finding has shown easy tiredness in physical activities even in pursuing the treatment procedure. These findings are consistent with a recent study on individuals with long COVID symptoms. Participants reported that COVID-19 had a profound impact on all aspects of their life and made them inactive compared to pre-COVID-19 normal (37). Moreover, another study showed adverse effects of long COVID symptoms on the ability of participants to engage in exercise, job duties, self-care, and housework that made them feel fatigued (38). As many studies have pointed out the significant role of activities (e.g., housework, gardening) in improving the mental health well-being of long haulers (39), this inactivity could negatively impact the mental health of COVID-19 long haulers. While some studies mentioned that COVID-19 negatively affects body image (40), our findings revealed that these changes in the body images were rooted in their physical limitations.

The economic losses to women are indirect consequences of the COVID-19 pandemic (41). The COVID-19 effect on the economy could emerge in different forms of a female long hauler’s life. Our results showed that unemployment and insecure financial status are part of the detrimental effects of COVID-19. Previous studies suggested that many women in the informal sector have limited access to benefits like social security, and widespread unemployment will cause long-term effects on their economic independence and security (42). Also, as women’s return to employment is often hindered when jobs are scarce, COVID-19 may worsen existing gender disparities over time (43). In addition, for women, job insecurity due to the coronavirus is another problem. Many studies demonstrate that women took on primary family responsibilities regardless of employment status when schools were closed, and family members were at home (44). Likewise, in our study, participants mentioned that they were laid off or forced to quit their jobs to attend family matters during the pandemic. Women’s higher family responsibilities make them more vulnerable to economic shocks than men (41). In this regard, our study participants reported that precarious economic conditions prevented them from paying for health care and medical treatment. These financial pressures, directly and indirectly, affected their recovery process.

This study found that conflict in social roles, particularly between work and family roles, was one of the outstanding issues experienced by female COVID-19 long haulers. Work-family conflict occurs when pressures from work and family roles cannot be adjusted in the right way (45). Therefore, employees who could not balance their time and energy between family and work have suffered from work-family conflict (46). Similar to our results, previous studies have shown that work-family conflict has a significant impact on the performance of female employees and brings them distress and exhaustion (46). During the COVID-19 pandemic, women have engaged in more household and caregiving responsibilities (47), culminating in raising multiple role conflicts (47, 48). As reported in our study, females experience job changes during COVID-19. These job changes put them in a difficult situation to choose between their health and the need to earn wages for paying living costs (47) and result in significant suffering for these females (49).

Our findings suggest that the attitudes of employers during the pandemic could be a critical factor on role conflict. Consistent with a study by Kong and Belkin (50), we found that employees feel neglected by their employers as a common experience of the pandemic. Considering the challenging situations of individuals in the pandemic, they anticipate attention and care from their employers (51). They want their employers to provide them a place where they feel seen and heard (52). Thus, as mentioned by our study’s participants, support from employers and coworkers in the workplace may help female employees cope better with difficulties related to role conflict at work (53-55) and can assist them to handle family and work roles more efficiently (56, 57).

The COVID-19 pandemic has also changed people’s social relationships and the way they communicate with each other. Algeri, Saladino (58) stated that the lockdown was characterized by changes to daily lifestyle, such as increasing time at home and decreasing social distance through digital devices. Some literature also suggested that elevated levels of social isolation and increased time spent at home lead to negative emotions (59-62). In our study, some participants stated that increased time spent at home and changes in time spent with family members made them feel irritable and stressed and affected their relationships with their family members. Participants who shared experiences of social isolation mentioned that the lack of understanding of the public about the persistent symptoms of long haulers and the disagreement of their friends about vaccination led to limitations in the development of their social relationships. Moreover, participants emphasized that only those who had similar experiences can understand them. Not everyone understands the difficulties experienced by long haulers, which leaves long haulers without timely and on-point help when they need advice and support the most.

Although social isolation affected the maintenance and development of social relationships among long haulers, this study found that social media use provided a novel way for long haulers to communicate. In support of previous research on social media use, the current findings focus on the advantages of social media use (64, 65), and that digital and social media and online platforms play a significant role in how people experience the threat of COVID-19 (66, 67). Based on the reasons such as social isolation and negative social response of persistent symptoms, long haulers tended to join peer support groups via online platforms and social media to communicate and share their experiences in order to gain social acceptance and improve their understanding of COVID-19 related information. Given that the pandemic has not yet ended, and people still face the “social distancing” policy, we need to pay attention to long haulers’ needs for social support from peers and be mindful of online intervention delivery.

According to our results, the social stigma associated with COVID-19 is one factor that adversely affects long haulers’ social lives. Imran, Afzal (68) showed that COVID-19 associated stigma might result in loss of trust or respect, rejection, and stigmatizing behaviors. In our study, participants reported that their social life have been impacted by being labeled, treated differently, or stigmatized during COVID-19. Likewise, other studies have shown that people feel a deep sense of exclusion due to stigma (69), which causes mental disorders (70). Furthermore, social stigma affects women’s health by being a prominent barrier to diagnostic and treatment procedures for improving mental health problems such as anxiety and depression (71). As in another study evaluating perceived stigma among patients with COVID-19 after recovery, those who scored higher on PTSD, anxiety, and depression reported more fatigue and stigma (72). Our study suggested that internalizing stigma, blaming themselves, and fear of being judged by other people exacerbate their psychological and physical burdens. The findings are consistent with other studies, which showed that people who have suffered from COVID-19 symptoms fear being judged by others (73), delay seeking health care, and being isolate (74).

Our study is limited by its generalizability. The small sample size of the interviews (n□=□15) did not allow for stratification by key demographic variables to assess the potential influence of race or ethnicity on their social life during long COVID. However, such qualitative research, typically based on small samples, may seek to provide trustworthiness and sufficient context to a greater extent to allow readers to make their own transferability judgement (75). Moreover, this study is part of an online health intervention among long haulers and recruited potential participants from Facebook groups, so they are a group interested in online intervention and have access to social media. Therefore, the participants may not be representative of the general population. Like all other qualitative research, our study may also be influenced by researchers’ subjective bias during the process of discussion guide development, transcription coding, and results interpretation. Finally, although two researchers conducted the coding independently, we did not calculate the intercoder agreement because they resolved all disagreements in the final coding through discussion.

## CONCLUSION

This study provides insights into the impact of long COVID on various aspects of social life among female long haulers. The persistent symptoms not only change their physical health but also lead to conflicts between their social roles. Findings highlight the need for more tailored social life-related advice and social support for long haulers. Future research needs to examine the impact of long COVID on the social life of other groups (e.g., men, people with disabilities, elderly, and immigrants), who may experience other types of social role conflict. Another pivotal subject we suggest based on this study is to explore in-depth new ways of online communication. Accordingly, this could pave the way for designing novel applications or providing various coping strategies for other pandemics. Regarding this issue, another study on coping strategies, especially among female long haulers, could be beneficial to provide information for different provider groups (e.g., policymakers, health care providers).

## Data Availability

All data produced in the present study are available upon reasonable request to the authors.

## DECLARATIONS

### Ethics approval and consent to participate

This study was reviewed and approved by the University of South Carolina Institutional Review Board (Pro00109358)

### Consent for publication

Not applicable.

### Availability of data and materials

The datasets used and analyzed during the current study are available from the corresponding author on reasonable request.

Competing interests

### There is no conflict of interest to report

#### Funding

Research reported in this publication was supported by the National Institutes of Health under Award #R01AI127203-5S1.

### Authors’ contributions

SQ and CY contributed to the study conception and design. Material preparation and data analysis were performed by AA and RZ. The first draft of the manuscript was written by AA and RZ, with ST and SQ also made major contribution in manuscript writing. All authors reviewed and revised the manuscript. All authors read and approved the final manuscript.

## Acknowledgements

Research reported in this publication was supported by the National Institutes of Health under Award #R01AI127203-5S1. The content is solely the responsibility of the authors and does not necessarily represent the official views of the National Institutes of Health.

